# Adaptation of the Walk ‘n Watch intervention for UK Community Stroke Rehabilitation: A Structured Adaptation Process

**DOI:** 10.64898/2026.05.01.26352175

**Authors:** Suzanne Ackerley, Sue Peters, Janice Eng, Stanley H Hung, Samantha Hancock, Cavan Smith, Nicola Keenan, Phil Woodford, Louise A Connell

**Affiliations:** Faculty of Health & Medicine, Health Innovation Campus, Lancaster University, UK; Rakehead Rehabilitation Centre, Burnley General Teaching Hospital, East Lancashire Hospitals NHS Trust, UK; School of Physical Therapy, Faculty of Health Sciences, Western University, ON, Canada; Gray Centre for Mobility and Activity, Parkwood Institute, ON, Canada; Department of Physical Therapy, University of British Columbia, BC, Canada; Centre for Aging Smart, Vancouver Coastal Health Institute, BC, Canada; Stroke Therapy Service, Pendle Community Hospital, East Lancashire Hospitals NHS Trust, UK; Lancashire and South Cumbria Integrated Stroke and Neuro Delivery Network, Lancashire, UK; Experienced Communications and Engagement professional and patient advocate, Lancashire, UK

**Keywords:** stroke, rehabilitation, mobility, gait, ADAPT guidance, implementation science, intervention development

## Abstract

**Background:** Walk ‘n Watch (WnW) is a structured, progressive walking exercise intervention developed for Canadian inpatient stroke rehabilitation. Although its mechanisms align with UK guidance for intensive walking therapy, stroke rehabilitation in the UK is delivered predominantly in the community. This change in service context has implications for safety, feasibility, and fidelity, necessitating structured pre-implementation intervention adaptation to support delivery.

**Methods:** A prospective adaptation process used ADAPT guidance. A multidisciplinary coalition and learning collaborative (UK clinicians, clinical- academics, people with lived experience, and Canadian WnW developers) participated in stakeholder co-production activities. Informed by ADAPT steps 1-2, co-production focused on rationale, core components, contextual mapping and planning adaptations. Discussions were analysed through rapid deductive mapping using Consolidated Framework for Implementation Research (CFIR) domains. Candidate fidelity-consistent adaptations were refined by the learning collaborative. Conceptual outputs of the process were synthesised.

**Results:** Three intervention core components were confirmed: 1) prioritised, high-volume, weight-bearing walking-related activities at moderate effort; 2) structured progression of steps based on performance on a walking test (e.g. Six-Minute Walk Test); 3) objective monitoring of steps and cardiovascular intensity. Several contextual determinants across CFIR domains were likely to influence UK community implementation. Fidelity-consistent modifications to the adaptable periphery were specified across four areas: 1) therapy & practice, 2) environment & safety, 3) monitoring & feedback, and 4) workflow & documentation. Adaptations included hybrid supervision, planned out-of-session practice, and monitoring using validated proxies. A WnW Adaptation Model was produced.

**Conclusions:** This paper provides a transparent pre-implementation adaptation of WnW for delivery within UK community stroke rehabilitation. Anchoring adaptations to intervention mechanisms and principles through co-production and implementation science frameworks, this work establishes a foundation for piloting and hybrid effectiveness–implementation evaluation. The WnW Adaptation Model offers support for future implementation efforts. Discussion positions adaptation as a pragmatic means for applying optimisation principles.

**PLAIN LANGUAGE TITLE:** Adapting the Walk ‘n Watch walking exercise programme for home-based stroke rehabilitation in the UK: A structured step-by-step process

**PLAIN LANGUAGE SUMMARY:** *Background:* Walk ‘n Watch (WnW) is a structured exercise programme that helps people improve their walking. It was originally developed for people recovering from stroke in hospital in Canada. While the approach fits well with United Kingdom (UK) recommendations for intensive therapy, stroke rehabilitation in the UK often takes place at home. Because of this difference, WnW needs careful adaptation for safe and effective delivery.

*Methods:* Published ADAPT guidance was used to adapt WnW. UK therapists, researchers, people with stroke, and Canadian WnW developers undertook adaptation activities. Together, they identified which parts of WnW were essential, explored differences between the Canadian and UK settings, and planned changes. Discussions were reviewed using an established framework to develop adaptations that kept the most important parts of WnW intact (fidelity-consistent adaptations). The adaptation process was summarised.

*Results:* Three essential intervention parts were confirmed: 1) prioritised, high-volume, weight-bearing walking-related activities at moderate effort; 2) structured progression of steps based on performance on a walking test; 3) objective monitoring of steps and cardiovascular intensity. Several factors were likely to influence delivery in the UK community. Changes focused on four areas: 1) therapy & practice, 2) environment & safety, 3) monitoring & feedback, and 4) workflow & documentation. They included using both in-person and online sessions, planning safe between session practice, and using non-digital monitoring. A WnW Adaptation Model was produced.

*Conclusions:* This paper clearly describes the steps taken to adapt WnW for delivery in UK community stroke rehabilitation. By working closely with stroke experts and using established research frameworks, the adapted programme keeps the most important parts of WnW while allowing it to fit into real-life. The WnW Adaptation Model offers support for further testing and may assist others looking to adapt WnW. Discussion offers perspective on how adaptation aligns with optimising interventions.

## INTRODUCTION

Walking endurance, gait speed and outdoor mobility remain persistent challenges for stroke survivors, limiting community participation after discharge (Blennerhassett et al., 2018). To address these post-stroke challenges in the UK, national stroke guidance recommends intensive, task-specific walking practice sufficient to produce a cardiovascular training effect, combined with principles such as structured progression, feedback and goal-setting (*National Clinical Guideline for Stroke for the UK and Ireland*, 2023). Evidence from systematic reviews indicates that intensive rehabilitation and aerobic exercise is safe and feasible across the stroke severity spectrum and is associated with meaningful gains in fitness, mobility and balance (French et al., 2010; Lloyd et al., 2018; Luo et al., 2020; Saunders et al., 2020). However, despite this, functional walking and mobility practice is commonly delivered at low dose and intensity within inpatient stroke rehabilitation in England (Tyson et al., 2018), an issue that can be amplified in community-based services (Clark et al., 2023). Translating principles of high dose, high intensity walking training into routine clinical practice can be particularly challenging in community rehabilitation due to constraints related to service configuration, resourcing and care environments (Clark et al., 2023).

Walk ‘n Watch (WnW) is a structured, progressive walking exercise intervention that has operationalised principles of neuroplasticity including high-repetition, intensity, and task-specificity within Canadian adult inpatient stroke rehabilitation (Peters et al., 2025; Peters et al., 2023). Delivered over 4 weeks (or until discharge), WnW requires patients to complete a minimum of 30 min of walking-related activity per physical therapy session. Intensity is progressed based on heart rate and step count monitoring, with targets derived from a baseline Six-Minute Walk Test (6MWT). In a large Phase 3 pragmatic multi-site stepped-wedge cluster randomised controlled trial, published in Lancet Neurology, WnW produced clinically meaningful improvements in walking endurance, gait speed, balance and quality of life, with no serious adverse events (Peters et al., 2025). Gains in walking endurance were largely maintained at 12 months poststroke (https://neurorehab.med.ubc.ca/walk-n-watch/).

Implementation studies for WnW highlighted both facilitators (e.g. progression guides, objective monitoring, clear planned routines) and delivery challenges (e.g. wearable device reliability, competing demands) (Ackerley et al., 2026; Hung et al., 2025), underscoring the importance of balancing fidelity with feasibility.

Given the strong evidence supporting the effectiveness of WnW intervention in Canada, a key translational question is how WnW might be delivered effectively, safely and feasibly in the UK National Health Service (NHS) context. In Canada, WnW was delivered in structured inpatient rehabilitation units with access to therapy gym spaces, consistent equipment and daily 30–60-minute therapy windows that support monitored cardiovascular progression. In the UK, stroke rehabilitation is delivered predominantly in the community, within patients’ homes and their neighbourhood (*National service model for an integrated community stroke service*, 2022; *Stroke Sentinel National Audit Programme (SSNAP)*, 2025), characterised by more variable physical spaces, less predictable access to assistance, and different workforce and workflow configurations. Implementation of WnW represents a logical next step in translating accumulated evidence into routine practice but raises important questions about how walking-specific dose, intensity and progression can be operationalised safely and consistently in the home and community environment. While WnW has demonstrated effectiveness in Canadian inpatient stroke rehabilitation, direct transfer to UK community services cannot be assumed to be safe, feasible, or fidelity-consistent. Structured pre-implementation adaptation is therefore required to preserve intervention mechanisms and principles while achieving contextual fit.

The aim of this project was to adapt WnW using ADAPT guidance (Moore et al., 2021), to support safe, feasible and fidelity-consistent delivery within UK community stroke rehabilitation, a substantively different context. Consistent with a pre-implementation focus, the objectives of the paper were to report ADAPT Steps 1–2 undertaken to: (i) assess the rationale for selecting WnW for UK community rehabilitation; (ii) confirm the WnW intervention core components; (iii) map contextual similarities and differences between the Canadian inpatient trial and UK community stroke rehabilitation service contexts; and (iv) specify WnW-UK fidelity-consistent adaptations. Considering scalability from the outset, a further objective was to; v) provide a model to synthesise the conceptual outputs of the adaptation process to inform future implementation efforts.

## METHODS

### Design and Overall Approach

This project represents the early phase of a structured intervention adaptation process corresponding to Steps 1 and 2 of the ADAPT guidance (Moore et al., 2021). Adapting existing interventions to a new context is a recognised and efficient approach to intervention development, as outlined in the Medical Research Council (MRC) Framework for Developing and Evaluating Complex Interventions (Skivington et al., 2021). Our prospective approach was therefore underpinned by the MRC framework, and methodological developments (Connell et al., 2025), and informed by prior experience applying ADAPT (Ackerley et al., 2025). Intervention adaptation described with ADAPT provided the procedural structure; the Consolidated Framework for Implementation Research (CFIR) guided contextual enquiry; and organisation of stakeholder engagement was informed by selected strategies from the Expert Recommendations for Implementing Change (ERIC) compilation (e.g. build a coalition; create a learning collaborative). Co-production activities included a series of stakeholder engagement activities, primarily workshops supplemented with consultations and iterative feedback (see Supplemental file 1), which together informed adaptation decisions rather than constituting formal qualitative data collection. Discussions were audio-video recorded or documented through field notes and analysed by the adaptation team through rapid deductive mapping using CFIR domains. The GUIDance for the rEporting of intervention Development (GUIDED) checklist was used (see Supplemental file 2) (Duncan et al., 2020).

### ADAPT Step 1: Assessing Rationale and Intervention–Context Fit

#### Purpose

A needs assessment was conducted to examine alignment between WnW’s mechanisms and principles against UK policy and service expectations, and to identify further contextual determinants relevant to optimising intervention-context fit for UK community-based stroke rehabilitation.

### Data Sources and Stakeholder Contributors

Information was collated from a range of documentary sources and stakeholder contributors. Intervention details and contextual information for the delivery conditions in the Canadian trial were drawn from the WnW published protocol, trial and implementation reports (Ackerley et al., 2026; Hung et al., 2025; Peters et al., 2025; Peters et al., 2023), as well as materials available on the WnW webpage (https://neurorehab.med.ubc.ca/walk-n-watch/). Contextual information on UK community stroke rehabilitation was drawn from national stroke guidelines and audit reports describing stroke pathways and service characteristics, and national policy documents outlining community-based rehabilitation workflow and workforce organisation, and wider healthcare priorities (*10 Year Health Plan for England: fit for the future*, 2025; *National Clinical Guideline for Stroke for the UK and Ireland*, 2023; *National service model for an integrated community stroke service*, 2022; *Stroke Sentinel National Audit Programme (SSNAP)*, 2025).

Documentary synthesis was combined with input from a WnW Stakeholder Coalition members, involving UK clinical-academics (n=3), NHS clinicians (n=12) and Canadian WnW developers (n=3), and WnW Learning Collaborative patient and public contributors (n=17), which was facilitated by established relationships. These stakeholder groups were considered essential in supporting intervention adaptation and future implementation efforts. The role and examples of their co-production activity are provided in Supplemental file 1. Inputs informed the structured needs assessment and did not constitute formal research data.

### Establishing Rationale

A clinician consultation workshop was held with UK coalition members to examine the need for and relevance of implementing WnW in the UK. This workshop also contributed to early contextual assessment and identification of anticipated barriers and facilitators (see below). Discussions focused on outer setting constructs related to guidelines and policy considerations including stroke guidelines and expectations for community-based stroke services, and whether differences between the service setting (i.e. inpatient vs. community) necessitated adaptation of the intervention.

### Identification of Intervention Core Components

Through iterative discussion via collaborative workshops and ongoing communication, UK clinical-academics and Canadian WnW developers confirmed the non-negotiable core components required to preserve WnW’s mechanisms of action, drawing on the original intervention logic model, trial experience, and outcomes. Screening for WnW intervention eligibility and safety was determined to be a fixed structural element, rather than an intervention core component, and was considered within the contextual assessment.

### Contextual Assessment

A structured comparison of context between the Canadian inpatient WnW trial and UK community stroke rehabilitation services was conducted across workshops. Contextual information was mapped using the CFIR to surface relevant contextual determinants, focusing on:

- Intervention characteristics: core and peripheral intervention components including timing, targets and technology
- Individuals characteristics: patient profile, carer involvement, therapist role
- Inner setting: physical environment, workflow and workforce patterns, resource

Implementation process was not the focus of this phase. The identification and specification of implementation activities and strategies to support enactment of WnW in the UK will be undertaken in later adaptation steps.

### Barrier/Facilitator Exploration

During consultation workshops (clinician, patient) and additional consultations, UK multidisciplinary clinicians, patients and their family members, discussed anticipated barriers and facilitators aligned with the contextual determinants. These insights informed areas for consideration when modifying the adaptable components (henceforth adaptable periphery) during Step 2 and were not treated as empirical data.

### ADAPT Step 2: Early Adaptation Planning

#### Purpose

To generate fidelity-consistent adaptations to enable safe, feasible delivery of WnW in UK community stroke rehabilitation.

### Stakeholder contributors

A WnW-UK Learning Collaborative supported shared decision-making and co-production of adaptations. A WnW-UK adaptation team jointly led adaptation specification and comprised UK clinical-academics (n=3), NHS community stroke physiotherapists (n=3), and a person with lived experience of stroke (n=1). Further learning collaborative members included wider NHS community and inpatient stroke physiotherapists and assistant practitioners (n=14), and patient and public contributors (n=17). Role and examples of co-production activity are provided in Supplemental file 1.

### Fidelity Criteria and Decision Rules

Core-components were non-negotiable and adaptations for context were retained only when the intervention core components remained intact.

### Planning Fidelity-Consistent Adaptations

The WnW-UK Adaptation Team participated in two collaborative adaptation workshops to map the adaptable periphery against Step 1 contextual determinants while maintaining the core components. Candidate fidelity-consistent adaptations were generated based on contextual assessment and barrier/facilitator exploration. Across two consultation workshops (patient, therapy staff), learning collaborative members subsequently refined candidate adaptations.

### Synthesising the early Adaptation Process

To strengthen fidelity and scalability for future implementation efforts, the conceptual outputs of ADAPT step 1 - 2 were synthesised into a WnW Adaptation Model. The model distinguishes non-negotiable intervention core components from areas for consideration when specifying fidelity-consistent adaptations to the adaptable periphery. Within this model, CFIR informed the organisation of contextual determinants and their linkage to corresponding fidelity-consistent adaptations key areas.

### Positionality Statement

The author team comprises UK clinical-academics, NHS clinicians, a UK person with lived experience of stroke, and Canadian WnW developers. Two authors (SA, LAC) are clinical-academics embedded within UK community stroke services, who collaborated on the Canadian WnW trial but were independent of the management team. Three authors (SP, SHH, JJE) are clinician-scientists from Canada who led the development and trial of WnW. This positioning provided valuable insight across both UK and Canadian contexts but also introduced potential interpretive bias. Three authors (SH, CS, NK) are UK NHS physiotherapists who contributed in-depth knowledge of the UK NHS community stroke services and helped ensure that adaptations were grounded in real-world service delivery. One further author (PW) is a person with lived experience of stroke, who contributed valuable experiential insight into the practicality and acceptability of the proposed adaptations. To mitigate potential bias, adaptation decisions were informed by multidisciplinary clinician input, early patient and public involvement, and structured use of implementation science guidance and frameworks to enhance transparency and reduce individual influence.

### Governance and Ethics

Activities in this early phase adaptation comprised stakeholder consultation and preparatory co-production; no patient intervention or formal data collection occurred. Accordingly, this project was classified as service-level engagement/pre-study development and did not require research ethics approval. Patient and public contributors provided experiential input; they were not research participants but regarded as members of the WnW-UK Learning Collaborative.

## RESULTS

### ADAPT Step 1: Rationale, Core Components and Contextual Determinants

#### Rationale for UK Adaptation

UK stakeholders agreed that WnW’s mechanisms and principles strongly aligned with national stroke guidelines and policy. The intervention reflects recommendations for intensive, repetitive, task-specific mobility and cardiovascular rehabilitation (*National Clinical Guideline for Stroke for the UK and Ireland*, 2023). They acknowledged an ambition by UK clinicians to provide high-intensity therapy, but current efforts were fragmented and siloed. WnW’s principles, together with its strong supporting evidence base, closely mirrored the expectations for personalised, responsive, high-quality rehabilitation outlined in UK community stroke service policy (*National service model for an integrated community stroke service*, 2022). The intervention’s encouragement of carer involvement, and its potential to promote self-management and community participation, were seen to strengthen alignment with policy intentions. Stakeholders also noted alignment with wider national priorities for digitally enabled, prevention-supportive, and community-based healthcare (*10 Year Health Plan for England: fit for the future*, 2025). Collectively, stakeholders felt that WnW could help support and standardise efforts to enact and sustain recommendations set out in UK national guidelines and policy.

However, stakeholders recognised that contextual differences in setting, in particular the transition to delivery in the community, would necessitate modification of the intervention. Community rehabilitation services typically provide shorter and less frequent supervised therapy sessions, operate under more variable delivery conditions, have less specialised equipment, and have diverse workflow and workforce patterns. These factors justified the need for structured pre-implementation adaptation of WnW to preserve fidelity while achieving safety and feasibility in the UK context.

### Intervention Core Components

Three intervention core components essential to WnW’s mechanisms of action were confirmed as non-negotiable:

1. Prioritised*, high-volume, weight-bearing walking-related activities at moderate effort
2. Structured progression of steps based on performance on a walking test (e.g. 6MWT)
3. Objective monitoring of steps and cardiovascular intensity

*’Prioritised’ refers to walking-related intervention activities being completed before other therapeutic activity.

Although the identification of WnW core components involved stakeholder discussion, these components were not generated de novo or by consensus alone. Rather, they reflect mechanisms intentionally embedded within the original intervention design and trial logic, grounded in established principles of neuroplasticity and exercise physiology (Klassen et al., 2020). High-volume, task-specific walking practice targets activity-dependent motor relearning; structured progression of dose and cardiovascular intensity reflects principles of progressive overload; and objective monitoring supports accurate delivery of cardiovascular training stimuli (Billinger et al., 2014; French et al., 2010; Klassen et al., 2017; Maier et al., 2019). The role of the adaptation process was therefore to confirm and protec**t** these mechanism-anchored components when transferring WnW to a new context, rather than to redefine the intervention content.

### Contextual Determinants Shaping UK Community Delivery

Structured contextual mapping using the CFIR demonstrated several determinants likely to influence delivery in the UK community setting. Table 1 provides a side-by-side comparison of the contextual determinants relevant to WnW delivery for the Canadian WnW inpatient trial and UK community stroke rehabilitation services, with key differences and similarities highlighted.

**Table 1.** Comparison of contextual determinants relevant to Walk ‘n Watch (WnW) delivery: Canadian inpatient trial vs UK community stroke rehabilitation services.

| Domain | Canadian Walk 'n Watch inpatient trial | UK Community stroke rehabilitation services | Key similarities and differences |
| --- | --- | --- | --- |
| **Intervention** | 4-weeks therapy prioritising, high-volume, walking-related activities at moderate effort. | Typically therapy is provided for up to 6-months. May include walking-related activities, but not necessarily prioritised, high-volume or at moderate effort. | **Adequate time window in service for intervention.**<br>**Walking-related activities are not necessarily prioritised, high-volume or moderate effort.** |
|  | 30-60 min supervised physiotherapy sessions, 5 days per week. Minimum 2-weeks (i.e. 10 sessions). Out-of-session practice encouraged if safe. | Therapy dose varies by patient and time post-stroke, typically 30-45min supervised physiotherapy sessions, 1-5 days per week. Total number of sessions variable. Out-of-session practice commonly prescribed if safe. | **Supervised therapy sessions shorter, typically less frequent.**<br>**Out-of-session practice commonly prescribed if safe.** |
|  | Step target set via 6MWT. | 6MWT not routinely used, limited space in homes. | **6MWT not routinely used, limited space.** |
|  | Heart rate reserve (HRR) target zone set via resting heart rate measures at baseline. | Resting heart rate usually measured on initial assessment, HRR typically not calculated. | **Resting heart rate usually measured at initial assessment, HRR not typically calculated.** |
|  | Monitoring tools for steps (Fitbit) and heart rate (HR) (Garmin Forerunner 235 watch) provided. Proxy measures sometimes used e.g. distance, Borg RPE, Talk test. | Monitoring tools for steps and HR not routinely provided or used. Mixed service and patient access to monitoring tools (i.e. wearables). Proxy measures sometimes used e.g. distance, Borg RPE, Talk test. | **Patients' steps and cardiovascular intensity not routinely monitored by a tool.**<br>**Proxy measures sometimes used.** |
|  | Physiotherapists recorded steps, minutes walking-related activity, and average HR for each session. Out-of-session practice not collected. | Physiotherapists complete clinical notes for each session, but steps and HR rarely recorded. Total minutes of motor function therapy recorded for national stroke audit but not specific to walking-related activity. Out-of-session practice not routinely captured. | **Step and HR rarely recorded daily.**<br>**Minutes specific to walking-related activity not routinely recorded.**<br>**Out-of-session practice data not routinely captured.** |
|  | Patients received an information booklet summarising intervention targets and progression plan. | Patients do not routinely receive written information on step or HR target progression. | **Patients do not routinely receive written information on step or HR target progression.** |
| **Individuals: Patients** | Medically stable adults (≥19 years) receiving rehabilitation ≤12 weeks after confirmed ischaemic or haemorrhagic stroke and:<br>- have a goal of walking<br>- able to walk ≥5 steps with max assistance x1<br>- able to understand and follow instructions for participation<br>- did not have any exclusion criteria<br>- confirmed safe for exercise using safety screening including BP, HR checks and sub-maximal exercise screen (6MWT)<br>[(http://neurorehab.med.ubc.ca/walk-n-watch/)](http://neurorehab.med.ubc.ca/walk-n-watch/) | Medically stable adults (≥18 years) receiving rehabilitation after confirmed ischaemic or haemorrhagic stroke with:<br>- variable; goals, mobility, communication and cognition, medical conditions | **Similar adult stroke rehabilitation population, typically medically stable.**<br>**Need to identify patients meeting additional inclusion and exclusion criteria.**<br>**Need to standardise process for specific safety screening.** |
|  | Median 22 days post-stroke (IQR 15-36). | Starts median 9 days post-stroke, median 42 days in service (IQR 23-76). | **Similar time window post-stroke.** |
|  | - 57% moderate-severe disability (Modified Rankin Scale (mRS) 4/5)<br>- Average walking speed 0.54m/s (SD 0.29)<br>- Median age 67y (IQR 60-77)<br>- 65% male, 92% white | - 17% mod-severe disability (4/5); 36% mod (mRS 3/5), 30% slight disability (mRS 2/5)<br>- Average walking speed variable, some may be unable to walk (hoist transfer) or require assistance<br>- Median age 75y (IQR 64-82)<br>- 56% male, ~90% white* | **More heterogenous disability, variable walking ability (hoist – full community ambulator), older median age.** |
|  | Variable confidence with mobility. | Variable confidence with walking indoors and, in particular, outdoors - with concerns relating to lack of objects to hold, uneven surfaces, weather, fear of falling, fatigue. | **Variable confidence with mobility, particularly when walking outdoors.** |
|  | Variable digital literacy and confidence. | Variable digital literacy and confidence. | **Variable digital readiness.** |
|  | Carer (family/friend/other) support where possible. | Carer (family/friend/other) support where possible. | **Carer involvement may be possible.** |
| **Individuals: Therapists** | Inpatient physiotherapists and therapy assistants provided walking-related therapy. | Community physiotherapists and assistant workforce provide walking-related therapy. | **Physiotherapists and assistant workforce provide walking-related therapy.** |
|  | Initial anxiety providing intensive exercise but confidence grew in environment with immediate assistance and peer-support. | Often low confidence providing intensive therapy in the home/community setting due to safety, space and time constraints, varied assistance. | **Often low confidence providing intensive therapy given home/community setting due to safety, space and time constraints, varied assistance.** |
|  | Variable digital literacy and familiarity with step and HR monitoring. | Variable digital literacy and familiarity with step and HR monitoring. | **Variable digital readiness.** |
| **Setting** | Single-site inpatient units and teams. | Distributed community teams visiting homes and community locations. | **Distributed community teams travelling to home and community locations.** |
|  | Often flexible session scheduling. | Pre-booked session scheduling with time constraints, inconsistent workflow. | **Pre-booked sessions with time constraints, inconsistent workflow.** |
|  | Controlled indoor space (e.g. gym, corridors), some access to outdoor space. | Less predictable indoor space (e.g. small, varied flooring, clutter) and outdoor space (e.g. varied terrains including surfaces and incline). | **Less predictable indoor and outdoor spaces.** |
|  | Access to mobility-related equipment including walking aids, assistive devices, treadmills, body weight support, bungee, weighted vests. | Typically, access to basic mobility-related equipment including walking aids and assistive devices. | **Typically, access to basic walking-related equipment only.** |
|  | Risk mitigation: Immediate access to assistance. | Varied access to assistance. | **Varied access to assistance.** |

\* Inpatient data, excludes missing reports.

### Screening for Eligibility and Safety

Inclusion criteria remain suitable overall; community service provides an appropriate population (adults with stroke, typically medically stable) however procedures to identify patients that meet the additional eligibility criteria and screening for safety, will need to be developed and align with local systems. The definition of assistance should be broad, to include family/carer assistance if safe.

### Areas for Consideration when Modifying the Adaptable Periphery

The contextual determinants identified indicated that WnW’s intervention core components could be preserved, but adaptable periphery required modification for optimising intervention-context fit. Assessment of the barriers and facilitators to implementation highlighted four key areas, aligned with contextual determinants, for consideration when specifying fidelity-consistent adaptations in Step 2:

- **Therapy & Practice:** Flexible supervised practice and safe out-of-session practice, may be needed due to shorter, less frequent supervised therapy sessions. Split sessions may help manage time constraints and address patients concerns about fatigue and fit with daily activities and appointments. Use of existing venues and groups may be beneficial to help achieve targets, but intervention mechanisms and principles need to be embedded. Carer involvement should be sought where available and safe.
- **Environment & Safety:** Graded exposure with planned progression of routines and routes may help accommodate constraints including limited space, variable outdoor terrain and weather conditions, and variable assistance. Creative solutions should be co-developed to help achieve and progress step and intensity targets where only basic mobility-related equipment is available, considering patient heterogeneity. Clear guidance is essential.
- **Monitoring & Feedback:** Provision of simple monitoring tools will be needed given inconsistent access to wearables among services and patients, with variable digital readiness. Proxy measures may support, especially if unable to monitor with a wearable safely or reliably during walking-related activity. Clear self-monitoring guidance is important, given the high likelihood of unsupervised practice. Therapists will require regular access to data and planned reviews with patients.
- **Workflow & Documentation:** Streamlined procedures and materials are needed for sharing

intervention information with patients, and recording steps, HR, and time spent on targeted walking-related activity, as these are not routinely recorded. Documentation needs to be able to be integrated within service clinical record systems to support communication across distributed teams. Procedures to facilitate adherence should be considered.

### ADAPT Step 2: Specification of Fidelity-Consistent Adaptations

Based on the Adapt Step 1, the WnW-UK Adaptation team generated a set of fidelity-consistent adaptations to support feasible implementation in UK community services. These were refined in consultation with the WnW-UK learning collaborative and are outlined in Table 2.

**Table 2:** Adaptable periphery mapped to the WnW Intervention Core Components for fidelity-consistent contextual fit in UK community stroke rehabilitation.

| **Core components (non-negotiable)** | **Adaptable periphery (fidelity-consistent)** |
| --- | --- |
| Prioritised, high-volume, weight-bearing walking-related activities at moderate effort. | Physiotherapist-led with assistant workforce involvement.<br>Involve carers (family/friend/other) when available and safe, ensuring appropriate education and support.<br>Up to five ≥30 mins supervised sessions per week until discharge from physiotherapy (at least 10 sessions within 4-weeks), allowing hybrid supervision (in-person/digital, physio/assistant sessions).<br>Prioritise the first 30 mins of supervised sessions to weight-bearing walking-related activities but consider split sessions if needed and safe (e.g. 2 x 15 mins – 15 mins supervised, 15 mins repeated practice out-of-session).<br>Facilitate out-of-session practice (if/when safe) to supplement or augment supervised sessions, providing clear guidance and promoting intentional application of the WnW mechanisms and principles (e.g. monitoring steps and cardiovascular intensity).<br>Facilitate use of existing venues (e.g. open gyms, leisure centres) and groups (e.g. community walking groups) and integrate into real-world tasks (e.g. errands), promoting intentional application of WnW.<br>Consider creative solutions for targeting appropriate effort, with everyday equipment (e.g. increasing speed, task complexity, hills, weighted rucksack; decreasing rests, assistance, aids etc).<br>Ensure all activities are tailored to the patient's ability. |
| Structured progression of steps based on performance on a walking test (e.g. 6MWT) | Complete a recommended walking test (e.g. 6MWT) to set step number target (if able). Gait speed from a validated walking test may provide an alternative means to set target.<br>Facilitate graded exposure to walking-related activity with planned progression of safe routines and routes (e.g. step/stair routines, micro-routes for small homes with use of corridor, outdoor routes with weather provisions), increasing repetitions, time, distance.<br>Provide clinicians and patients with simple written (and/or online) information materials; including explanation for how WnW works, supporting evidence. Patients should receive clear, individualised, targets and practice guidance. Guidance should stipulate safety for out-of-session practice including where (indoor/outdoor), level of supervision (supervised/unsupervised) and support (assistance/aid). |
|  | Develop streamlined procedures to ensure regularly scheduled check-ins (in-person/digital/telephone) between physiotherapist and patient to support intervention adherence and progression.<br>Prompts should be considered between supervised contacts to help maintain adherence. |
| Objective monitoring of steps and cardiovascular intensity | Calculate target heart rate zone (40-60% HRR) on initial session.<br>Provide patients with HR monitor and step counter.<br>*Requirements:*<br>- Useable and acceptable for heterogeneous patient use<br>- Patient display for self-monitoring steps and HR (if able)<br>- Ability for therapy staff to regularly collect data (minimum: in session, ideally: +out-of-session)<br>- Reasonable battery life before charging<br>Monitor using validated non-digital proxies (e.g. BORG RPE, talk test) for cardiovascular intensity in parallel with tools. Use this for recording daily out-of-session practice if use of a monitoring tool is not possible.<br>Provide patients with simple daily activity monitoring form (patient choice of paper-based or online) for capturing steps, HR and time spent on walking-related activity in sessions and during out-of-session practice, allowing for recordings from monitoring tool and validated proxies.<br>Educate carers to support monitoring if needed and possible.<br>Develop streamlined materials for community stroke physiotherapists and assistants to input clinical records relating to screening, baseline measures, targets and ongoing monitoring that integrate with clinical record systems and outline plans for data review and patient check-ins. |

### Synthesis: WnW Adaptation Model

Conceptual outputs from the adaptation process were synthesised into a WnW Adaptation Model (Figure 1).

**Figure 1.**
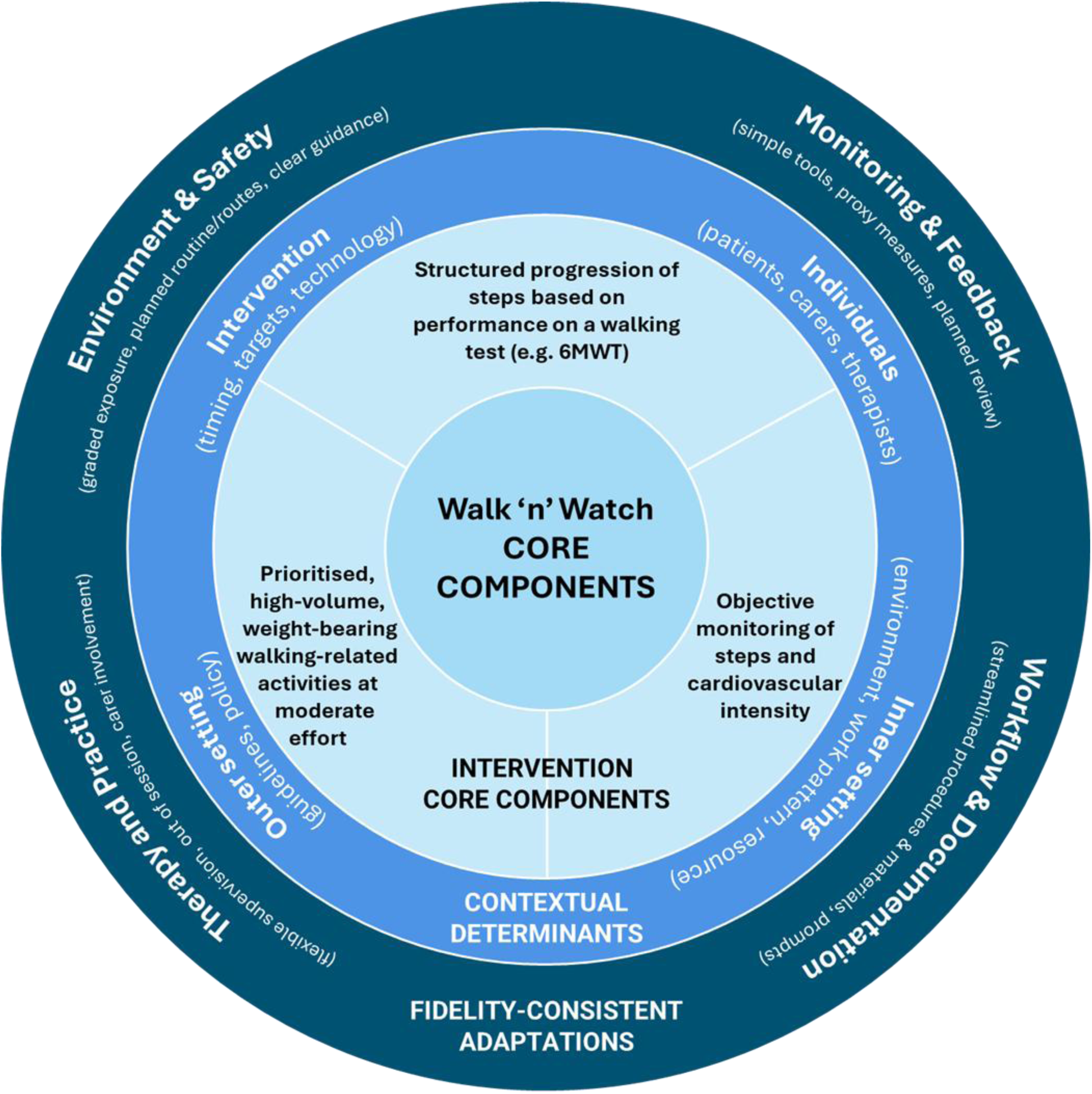
Walk ‘n Watch (WnW) Adaptation Model showing the synthesis between WnW’s intervention core components (centre), CFIR-informed contextual determinants (middle layer) and key areas for consideration when specifying fidelity-consistent adaptations to the adaptable periphery (outer layer).

## DISCUSSION

This paper provides a transparent account of structured, pre-implementation adaptation of the WnW intervention to support safe, feasible and fidelity-consistent delivery within UK community stroke rehabilitation. Using ADAPT guidance, and informed by CFIR and co-production with a wide range of relevant stakeholders, we confirmed rationale, non-negotiable intervention core components, mapped contextual determinants shaping delivery, and specified fidelity-consistent adaptations to the adaptable periphery. This project provides a mechanism-anchored foundation for subsequent piloting and hybrid effectiveness–implementation evaluation. The resulting WnW Adaptation Model is intended to support future implementation efforts, including aiding fidelity assessment and considering scalability.

### Why structured adaptation was required in a new service context

The WnW intervention has previously been iterated and evaluated within Canadian inpatient stroke rehabilitation through a large stepped-wedge implementation trial (Peters et al., 2025). The progressive walking exercise intervention was originally developed and tested in the explanatory ‘Determining Optimal post-Stroke Exercise’ (DOSE) trial (Klassen et al., 2020). Following demonstration of efficacy, refinements to both the intervention and its implementation strategies were undertaken to improve feasibility and delivery for real-world application, informed by associated qualitative findings (Connell et al., 2018; Janssen et al., 2020). Effectiveness of this iterated intervention was subsequently demonstrated in the Phase 3 cluster RCT, alongside a strong shift towards pragmatism (Ackerley et al., 2026; Peters et al., 2025).

For the present project, substantive contextual transfer across nations, systems, and services, warranted successive structured pre-implementation adaptation, rather than direct replication. Without this preparatory work, the intervention risked informal modification during early delivery, potentially compromising safety or eroding core components. Structured, co-produced, adaptation enabled contextual determinants to be explicitly identified and addressed in advance, supporting principled decision-making about what must be preserved and what could be modified to achieve safe and feasible delivery of WnW in UK community stroke rehabilitation. Intervention core components were deliberately bounded by established neuroplasticity and exercise physiology principles and reflected the original intervention logic and trial evidence, and adaptations were governed by decision rules to ensure they were not compromised in the transition to the new service context.

### Adaptation and optimisation as complementary processes

Intervention adaptation and optimisation can be presented as distinct or sequential processes, with adaptation framed as a response to contextual mismatch and optimisation framed as refinement of intervention components and/or implementation strategies in response to constraints (Guastaferro & Collins, 2021; Moore et al., 2021). However, particularly within implementation research, it has been highlighted that these processes have synergies (Guastaferro & Collins, 2021; Guastaferro et al., 2025). When interventions are transferred to new contexts, structured adaptation frequently contributes to optimisation for implementation by improving intervention–context fit and constraining unsafe or unplanned variation. Accordingly, we position our early-phase adaptation as complementary to the preparatory phase of optimisation.

In this project, the adaptation process focused on improving the conditions under which WnW’s core components could be enacted safely and consistently within the realities and constraints of UK community stroke rehabilitation. ADAPT guidance provided the most appropriate methodological framework for this purpose. By confirming the rationale and core components, and aligning intervention adaptations to reinforce these, this work aligns with the preparatory logic underpinning approaches such as Multiphase Optimisation STrategy (MOST) (Guastaferro & Collins, 2021; Guastaferro et al., 2025; Landoll et al., 2022). Our understanding appears congruent with how others have interpreted the early preparation phase of MOST as a formative stage for intervention adaptation (Whitesell et al., 2019). This project does not extend into the experimental comparison of alternative components or dose parameters undertaken in the specific optimisation phase of MOST. In line with an overarching approach underpinned by the MRC framework and developments (Connell et al., 2025; Skivington et al., 2021), adaptations were generated through early stakeholder engagement and actively considered economics, sustainment and scalability. Thus, the implementation science approach fits with the broader conceptualisation of intervention optimisation that emphasises balancing effectiveness with real-world implementation constraints (e.g. intervention EASE).

Importantly, adaptations avoided claims about optimising effectiveness but ensure that any subsequent optimisation or hybrid evaluation is conducted on an intervention that is sufficiently stabilised for the new context.

### Remaining uncertainties for later-phase adaptation, piloting and hybrid evaluation

As with all early-phase adaptation work, this project did not resolve all sources of uncertainty. Introducing an adapted intervention into a new context inevitably requires the tailoring of implementation strategies that support its delivery. In this project, we intentionally focused on adapting the intervention, rather than specifying or testing implementation strategies in detail. This reflects a staged method recognising that specification of implementation strategies is most meaningful once the intervention itself is sufficiently refined and stabilised for the new context. Explicitly separating intervention adaptation from implementation strategy optimisation reduces the risk of conflating intervention misfit with implementation failure during future evaluation.

Selection and evaluation of implementation strategies—such as training, supervision, workflow integration and monitoring processes—will be addressed in subsequent adaptation steps. In a previous study, ten implementation strategies were deemed successful in supporting WnW delivery in the Canadian trial, identified retrospectively from the perspectives of front-line therapists and managers using the CFIR-ERIC matching tool (Ackerley et al., 2026). These strategies provide a useful starting point for determining candidate implementation strategies which may be triangulated with strategy matching informed by the contextual determinants identified in the current project.

Key areas requiring empirical examination during piloting and hybrid evaluation include supervision configurations (e.g. in-person, hybrid and assistant-supported delivery), practical approaches to monitoring steps and cardiovascular intensity in routine practice, and the balance between supervised and out-of-session walking needed to achieve sufficient dose while managing risk. The resource implications and workflow impact of different delivery configurations also need further consideration. These uncertainties are deliberately structured rather than residual shortcomings. By making them explicit, this project provides a clear agenda for subsequent piloting, hybrid evaluation and, where appropriate, future optimisation of delivery configurations and implementation strategies.

### Preparing Walk ‘n Watch for hybrid effectiveness–implementation evaluation

A central ambition of this adaptation work was to prepare WnW for hybrid effectiveness–implementation evaluation in UK community stroke services. Hybrid designs seek to assess both intervention outcomes and implementation processes (Curran et al., 2012); however their interpretability depends on the intervention and accompanying implementation strategies being sufficiently well described and contextually appropriate at the point of testing. Undertaking further structured adaptation (ADAPT steps 3-4) to address the uncertainties described above in advance of hybrid evaluation will reduce avoidable risk that could otherwise obscure interpretation of both implementation and effectiveness findings.

Of note, the adaptation process highlighted heterogeneity within the target population. In particular, digital approaches to monitoring raise considerations relating to access, literacy and confidence. To mitigate the risk of digital exclusion, the adapted intervention specifies the use of validated non-digital proxy monitoring options alongside wearable technologies. Further piloting is required to examine how different monitoring approaches perform across diverse patient subgroups and whether additional tailoring is needed to ensure equitable access and engagement. Targeted evaluation within a hybrid trial may therefore be warranted.

### Strengths and limitations

This project was strengthened by its use of established adaptation guidance, implementation science framework (CFIR), and co-production with a wide range of stakeholders, including lived-experience input. Core intervention components and decision rules and adaptation processes undertaken were described transparently. The development and reporting of the WnW Adaptation Model provide a robust foundation for guiding local site adaptations and supporting fidelity assessment within a hybrid effectiveness-implementation trial, and for considered scaling to other contexts, contingent on evaluation.

However, this project does not provide empirical data on the effectiveness of the adapted intervention and is specific to UK community stroke rehabilitation. Implementation strategies were not specified at this stage.

These limitations reflect the project’s preparatory focus and will be addressed in subsequent adaptation, piloting and evaluation phases.

## CONCLUSION

Structured, co-produced adaptation of Walk ‘n Watch enabled preparation of the intervention for safe and feasible delivery in UK community stroke rehabilitation while preserving its core mechanisms and principles. By addressing contextual fit prior to evaluation, this work stabilises the intervention for the new context and provides a robust foundation for subsequent hybrid effectiveness–implementation research. The WnW Adaptation Model synthesised the adaptation process and is intended to support future fidelity assessment and scalability. More broadly, this project deliberates the intersection of adaptation and optimisation in implementation science when interventions with established effectiveness are transferred to new contexts. It illustrates the importance of revisiting optimisation assumptions when delivery conditions change and positions structured adaptation as a pragmatic mechanism for applying optimisation principles.

## ACKNOWLEDGEMENTS

The authors would like to thank the members of the WnW Stakeholder Coalition and WnW-UK Learning Collaborative.

## STATEMENTS AND DECLARATIONS

### Ethical Approval and Informed Statement

There are no human participants in this article and informed consent is not required.

### Data Availability Statement

Data sharing is not applicable to this article as no datasets were generated or analysed during the current project.

## Supplemental file 1

**Table S1:**
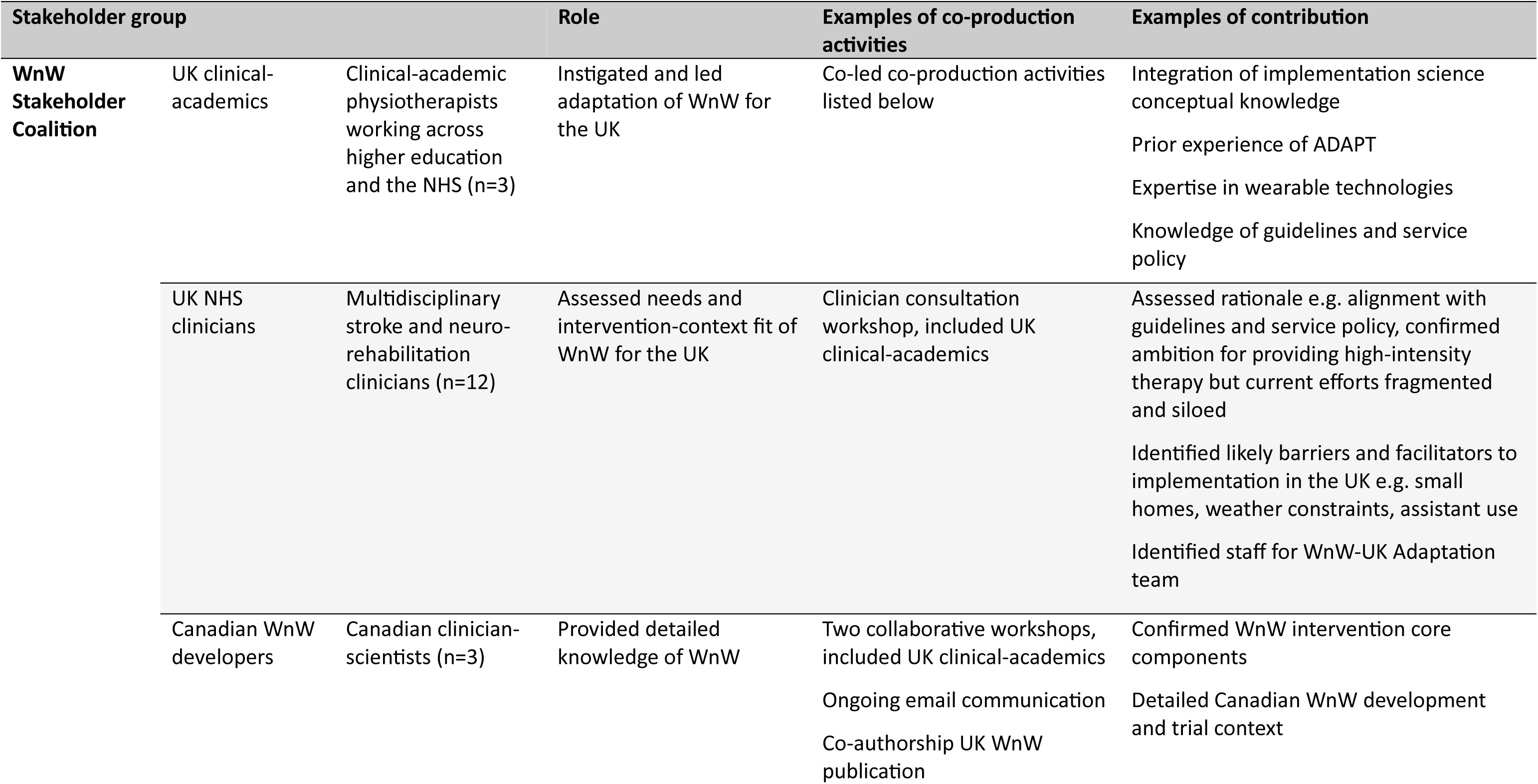
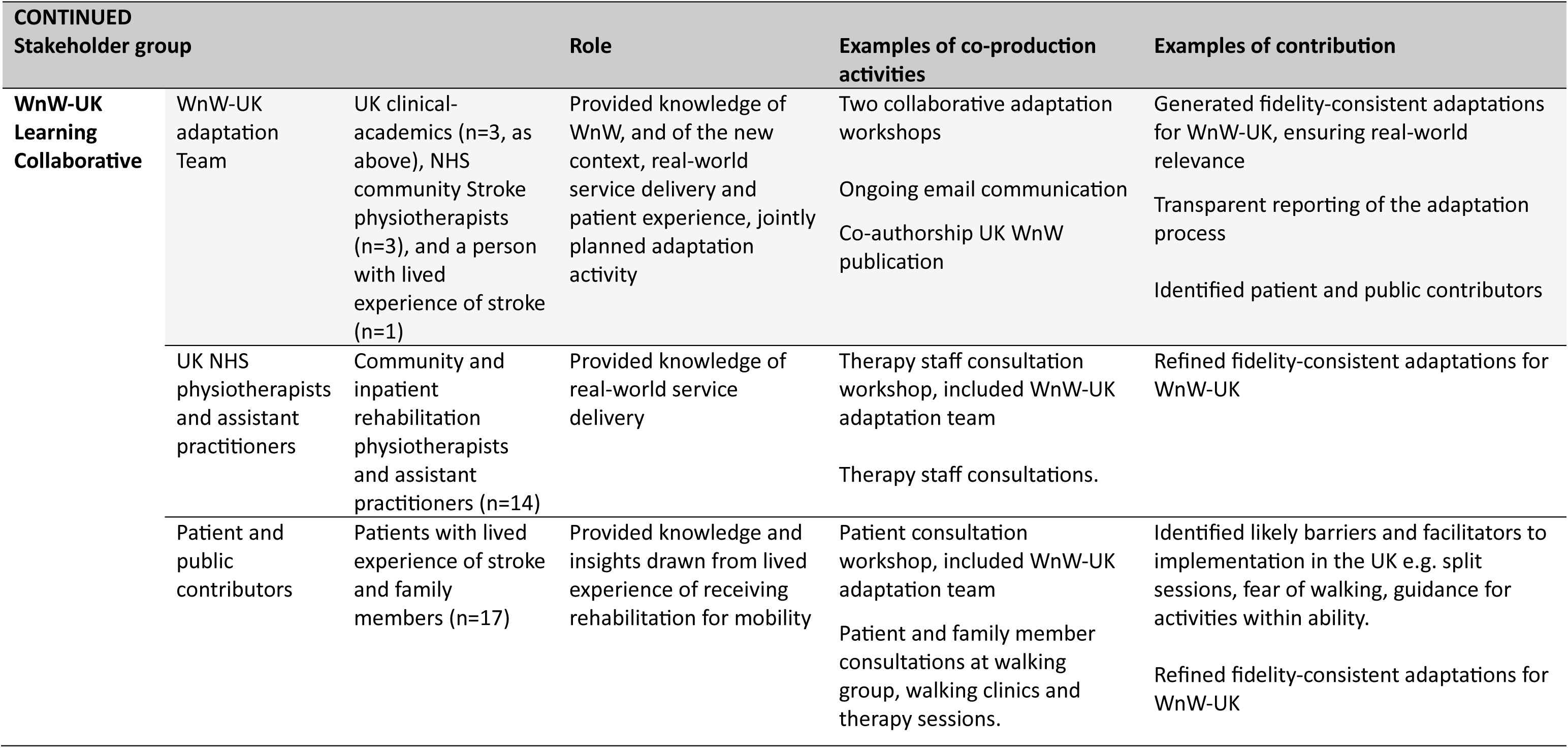
Stakeholder groups and co-production activities.

## Supplemental file 2

**Table S2:** GUIDED Checklist*. For Adaptation of the Canadian inpatient Walk ‘n Watch intervention for UK Community Stroke Rehabilitation: A Structured Adaptation Process (Ackerley et al., *submitted*)

| Item description | Page in manuscript where item located | Other* |
| --- | --- | --- |
| 1. Report the context for which the intervention was developed. | Table 1 |  |
| 2. Report the purpose of the intervention development process. | Page 7-9<br>Page 15-16 |  |
| 3. Report the target population for the intervention development process. | Table 1 |  |
| 4. Report how any published intervention development approach contributed to the intervention development process. | Page 9-10 |  |
| 5. Report how evidence from different sources informed the intervention development process. | Page 10-11 |  |
| 6. Report how/if published theory informed the intervention development process. | Page 9-10 |  |
| 7. Report any use of components from an existing intervention in the current intervention development process. | All |  |
| 8. Report any guiding principles, people or factors that were prioritised when making decisions during the intervention development process. | Pages 11-15 |  |
| 9. Report how stakeholders contributed to the intervention development process. | Pages 11-15<br>Supplemental file 1 |  |
| 10. Report how the intervention changed in content and format from the start of the intervention development process. | Table 2 |  |
| 11. Report any changes to interventions required or likely to be required for subgroups. | Pg 30 |  |
| 12. Report important uncertainties at the end of the intervention development process. | Pg 28-29 |  |
| 13. Follow TIDieR** guidance when describing the intervention. | Pg 7<br>Table 1<br>Table 2 |  |
| 14. Report the intervention development process in an open access format. | For submission to an open access peer-review journal, with pre-print made available |  |

\*Duncan, E., O'Cathain, A., Rousseau, N., Croot, L., Sworn, K., Turner, K. M., Yardley, L., & Hoddinott, P. (2020). Guidance for reporting intervention development studies in health research (GUIDED): an evidence-based consensus study. *BMJ open*, 10(4), e033516.
\*\*The Template for Intervention Description and Replication (TIDieR)-Rehab extension was used (Signal, N. et al.,2024), https://doi.org/10.1136/bmjopen-2024-084320)

